# Divergent transcriptomic profiles in depressed individuals with hyper- and hypophagia implicating inflammatory status

**DOI:** 10.1101/2023.09.25.23296077

**Authors:** Shu Dan, Julia Hall, Laura M. Holsen, Torsten Klengel

## Abstract

Major Depressive Disorder (MDD) is a heterogenous and etiologically complex disease encompassing a broad spectrum of psychopathology, presumably arising from distinct pathophysiological mechanisms. Divergent appetitive phenotypes including Hyperphagic MDD (characterized by an increased appetite) and Hypophagic MDD (characterized by a decrease in appetite) are important clinical characteristics that are closely related to comorbidities, including cardiometabolic disorders. Prior evidence supports the notion that hyperphagia is associated with atypical depression, decreased stress-hormone signaling, a pro-inflammatory status, hypersomnia, and poorer clinical outcomes. Yet, our understanding of the underlying mechanisms of Hyperphagic and Hypophagic MDD is limited, and knowledge of associated biological correlates of these endophenotypes remain fragmented. We performed an exploratory study on peripheral blood RNA profiling using bulk RNAseq in unmedicated individuals with Hyperphagic and Hypophagic MDD (n=8 and n=13, respectively) and discovered individual genes and gene pathways associated with appetitive phenotypes. In addition, we used the Maastricht Acute Stress Task to uncover stress-related transcriptomic profiles in Hyper- and Hypophagic MDD.

## Introduction

The phenotypic and etiologic heterogeneity in Major Depressive Disorder (MDD) remains a significant challenge toward establishing a better understanding of the underlying molecular mechanisms leading to MDD and in turn more effective and personalized treatments. Differences in appetite and weight are common among depressed individuals, with approximately half experiencing a loss of appetite and about a third of MDD patients exhibiting an increase in appetite^1^. An increased appetite along with weight gain is a feature of atypical depression whereas a decrease is commonly associated with melancholic depression. However, despite prior evidence suggesting that changes in appetite and weight are relevant long-term markers of divergent endophenotypes in depression, and although they can be easily assessed, these phenotypes remain understudied^2^.

Recent data highlight, in part, the neurobiological underpinnings of opposing appetite-related phenotypes of MDD. At the immune and hormone level, depressed individuals with increased appetite show associations with decreased HPA axis activation^1^, increased inflammation^3^, and insulin resistance^1^, while those with decreased appetite exhibit elevated ghrelin secretion in response to a meal^4^. At the neural systems level, there is initial evidence of hyperactivity in ventral striatum, pallidum, and putamen among Hyperphagic MDD subjects in response to food cues, and attenuated activation in the insula in Hypophagic MDD^5^. During rest, there is evidence of aberrant and divergent connectivity between nucleus accumbens, medial prefrontal cortex, insula, and orbitofrontal cortex in these groups compared to healthy controls^6,7^. Moreover, we and others have noted associations between activation to food cues and hormone (including ghrelin and cortisol) and immune (CRP, IL-1RA, IL-6) markers, in opposing directions for Hyper- and Hypophagic MDD^1,4,8^. Taken together, these findings converge to suggest distinct associations between immunometabolic and hormone markers and broad reward and interoceptive network aberrations that track with divergent appetitive phenotypes of MDD. However, few studies have examined the molecular basis underlying appetitive and stress responsivity in Hyper- and Hypophagic MDD, which could play a critical role in the development and maintenance of appetite increases and decreases in MDD.

Molecular analyses of individuals with Hyperphagic and Hypophagic MDD provide further evidence for genetic^9^ and transcriptional^10^ differences. An early transcriptomic study in n=1212 individuals (n=331 healthy controls, n=246 hyperphagia, and n=341 hypophagia) by de Kluiver et al revealed transcriptomic changes in line with an increased inflammatory status in Hyperphagic MDD compared to controls. This pro-inflammatory phenotype was strongly attenuated in Hypophagic MDD vs. controls. De Kluiver et al concluded that hyperphagia represents a distinct depressive subtype with partly differential pathophysiology regarding immune-metabolic processes that may benefit from more personalized anti-inflammatory treatment strategies. While de Kluiver emphasized the importance of the Hyperphagic subtype of MDD, we hypothesized that contrasting hyper- and hypophagia directly in an unbiased way using RNAseq would allow us to uncover additional transcriptional profiles that disentangle both phenotypes. In addition, considering the critical role of stress in appetitive function and in the pathophysiology of MDD, we introduced the Maastricht Acute Stress Test (MAST) which elicits a strong autonomic and glucocorticoid stress response, to uncover additional stress-related transcriptomic changes as a function of hyper- and hypophagia.

## Methods

### Participants

As part of a larger study on the relationship between stress and hormone function in MDD, adults with Major Depressive Disorder and healthy controls without current psychiatric disorders 21 and 45 years of age and with a BMI between 19 and 45 kg/m^2^ were recruited from the greater Boston area through online advertisements. From the larger sample of participants who completed all study components, RNA samples for the current analyses were collected on a subset of 21 individuals with Hyperphagic and Hypophagic MDD (8 Hyperphagic MDD [5 female]; 13 Hypophagic MDD [7 female]). Exclusion criteria included the history of substance abuse; current comorbid psychiatric disorders; current psychotropic medications use; history of neurological disease; endocrine disorders; diabetes; cardiovascular disease; current treatment with weight loss medications; glucocorticoids; steroids; current suicidal ideation; traumatic brain injury; for females, pregnancy or breastfeeding, and past amenorrhea greater than three months. All participants with MDD met diagnostic criteria as assessed by the Structured Clinical Interview for Diagnoses (SCID; DSM-IV-TR)^11^ administered by a trained clinician interviewer with over 20 years of experience. Participants with MDD were assigned into Hyperphagic MDD (e.g., increased appetite/weight gain) and Hypophagic MDD (e.g., decreased appetite/weight loss) groups based on their responses to appetite- and weight-related assessment in the mood disorders module of the SCID. Specifically, MDD group was assigned based on responses to criterion Question A3 under the “A. Mood Episodes” module: “(3) significant weight loss when not dieting, or weight gain (e.g., a change of more than 5% body weight in a month) or decrease or increase in appetite nearly every day for the past 2 weeks”. Participants who endorsed either weight loss OR decrease in appetite nearly every day for 2 weeks (A4) were categorized as Hypophagic MDD. Participants who endorsed either weight gain OR increase in appetite nearly every day for 2 weeks over the past month (A5) were categorized as Hyperphagic MDD. All participants provided written informed consent. All study procedures were approved by the Mass General Brigham Institutional Review Board.

### Procedures

Following a screening session to establish MDD status and obtain height, weight, anthropometric measurements, and hematocrit level (<34% in females and <37% in males), eligible participants underwent two main study visits (1 week apart) that were identical except one included a stress-inducing version (Stress) and the other included a non-stressful control version (No Stress) of the Maastricht Acute Stress Task (MAST)^12^. Visit order (Stress, No Stress) was randomized and counterbalanced across groups. Visits occurred between 0800 hrs and 1300 hrs following a 12-hour overnight fast. For females, all visits occurred during the follicular phase of the menstrual cycle. Upon arrival, participants completed the Beck Depression Inventory (BDI-II) to assess current depressive symptoms^13^; female participants underwent a pregnancy test. Next, a nurse placed an intravenous catheter (IV) with a saline lock for blood sampling. After the IV was placed, participants consumed a breakfast meal (within a 15-minute period to standardize rate of intake) designed to comprise 30% of their recommended daily caloric intake (see detailed description below). The T0 blood draw was completed immediately following the breakfast session. At this time, participants completed an assessment of their mood using an electronic visual analog scale (VAS) system^14^.

Participants then completed either the Stress or No-Stress version of the MAST. Participants were seated in front of a computer, and a water bath was positioned on the side of the arm that did not have the IV. Participants were then introduced to a female experimenter posing as a doctor who provided detailed instructions for the water and math task. This was followed by 10 minutes during which the participants alternated between five hand immersion trials (60 sec to 90 sec) and four arithmetic trials (30 sec to 90 sec). During the task, trial switching was prompted by a timed slideshow running on the computer (**Figure 1**).

**Figure 1.**
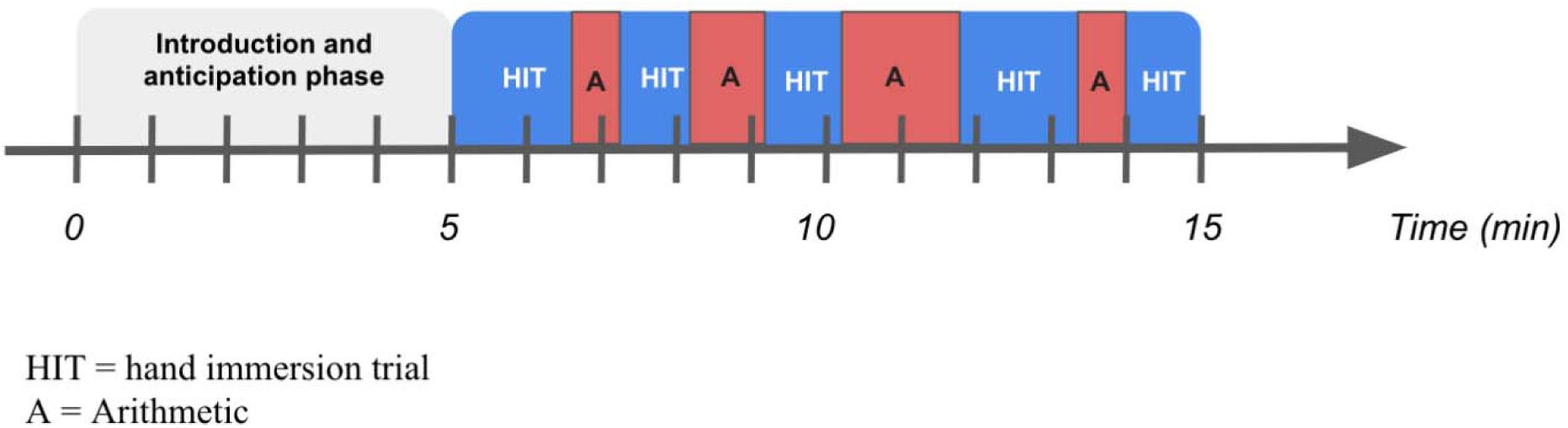
Schematic depiction of the Maastricht Acute Stress Task (MAST)

During the Stress visit, hand immersion trials required the participant to place their hand and wrist in cold water that was held between 0°C and 2°C. Arithmetic trials required counting backward as quickly and accurately as possible from 2,043 in intervals of 17. If a mistake was made, the experimenter instructed the participant to start again from 2,043. Participants were told their performance was being videotaped by a camera that was mounted to the computer screen. In reality, the camera was not recording. Following the final hand immersion trial, participants were told that their performance was poor and that they would need to repeat the task later during the visit; this manipulation was used to induce sustained levels of stress throughout the visit.

During the No-Stress visit, the water temperature was lukewarm (25°C – 37°C), and the math trials required counting up consecutively from 1 to 25 in steps of 1 at their own pace and starting over once they reached 25. There was no mention of videotaping, and the experimenter gave no feedback on their performance. The study staff member playing the role as doctor was consistent for each participant across Stress and No-Stress visits.

Approximately 85 minutes after the MAST (105 minutes since T0), a second sample of blood was drawn while participants completed a second mood assessment via the VAS system. At the end of each visit, participants were fully debriefed. During the No-Stress visit, participants completed a questionnaire packet including the trait portion of the State-Trait Anxiety Inventory (STAI)^15^, Perceived Stress Scale (PSS)^16^, and the Dutch Eating Behavior Questionnaire (DEBQ)^17^.

### Anthropometry

At the screening visit, height (cm) and weight (kg) were measured using a stadiometer. Those values were used to calculate body mass index (BMI [kg/m^2^]).

### Breakfast meal

At the start of each visit, participants ate a breakfast meal that was standardized to meet 30% of their daily caloric intake. Breakfast was prepared by Center for Clinical Investigation (CCI) dietary staff at Brigham and Women’s Hospital and varied according to each participant’s basal metabolic rate and physical activity level, measured by the Harris-Benedict equation with 18% calories from protein, 23% calories from fat, and 59% calories from carbohydrates^18^. Participants were encouraged to consume as much of the meal that they could in 15 minutes. After the 15-minute period, CCI dietary staff weighed the remaining food items to determine nutrient intake for total kcals and kcals per macronutrient. Planned versus consumed (PVC) kcals for total breakfast meal and then each individual macronutrient (protein, fat, and carbohydrate) was calculated by dividing the intake values by the planned standardized amounts.

### Blood sampling, hormone measurement, and RNA processing

Blood was sampled at three time points at each visit. Serum cortisol, acylated ghrelin, and RNA were collected at two time points: T0: After the breakfast meal/before the MAST and T105: 105 minutes after T0. Serum cortisol, alone, was collected at T20 as a manipulation check for the effect of the MAST.

For cortisol, after transferring to a tube containing pefabloc, serum was frozen until assayed. Serum samples for cortisol and ghrelin were stored at −80°C in plastic tubes containing a 10-mg/ml solution of PMSF (phenylmethanesulfonylfluoride) in methanol. Cortisol samples were assayed by LabCorp (Raritan, NJ) using electrochemiluminescence immunoassay (ECLIA) on a Roche Cobas analyzer; intra-assay CV 1.0-1.7%; inter-assay CV 1.4-2.2%.

Acylated ghrelin was collected in Ethylenediaminetetraacetic acid (EDTA) tubes, aliquoted with HCL, and then centrifuged. Ghrelin samples were assayed by the Brigham Research Assay Core (BRAC) using an enzyme immunoassay (ELISA; Millipore, St. Charles, MO; intra-assay CV 0.8-7.5%; inter-assay CV 3.9=12.9%).

Total RNA was collected in Tempus Blood RNA tubes (Thermo Fisher Scientific, Waltham, MA) incubated at room temperature for 2 hours and stored at −80°C according to the instructions of the manufacturer until further processing. Total RNA was extracted using the Norgen Total RNA Purification Kit (Cat. 17200, Norgen Biotek Corp, Thorold, ON, Canada) and quantified using a Qubit 2.0 Fluorometer (Thermo Fisher Scientific Inc., Waltham, MA). All samples were run on an Agilent 2100 Bioanalyzer (Agilent, Technologies, Santa Clara, CA) to determine RNA Integrity Number (RIN) and all samples passed QC with RIN values >6 (mean = 8.81, SD = 0.46).

### Behavioral measures

#### Beck Depression Inventory (BDI-II)

The BDI-II is a 21-item self-report questionnaire used to assess presence and severity of depressive symptoms^13^. Each item presents a characteristic (e.g., ‘Self-Dislike’) with four statements below it (labeled 0, 1, 2, and 3), with 0 representing minimal change in/severity of, and 3 representing maximal change in/severity of that characteristic. Participants were asked to indicate to what extent they experienced that characteristic in the past two weeks by choosing the statement they agreed with. Scores were calculated by summing the answers to each question. Higher scores indicate more severe depressive symptoms.

#### Mood Assessment

Ratings of sadness and tension were measured during the T0 and T105 blood draws using an electronic visual analog scale (VAS) system^14^. Participants were asked to rate how sad and how tense they felt by moving a slider on a linear scale that went from 0 to 100, with 0 being the least sad/tense they have ever felt and 100 being the saddest/tensest they have ever felt.

#### Dutch Eating Behavior Questionnaire (DEBQ)

During the No-Stress visit, participants completed the DEBQ, a 33-item self-report questionnaire that assesses three eating behavior domains: Restrained Eating (10 items), Emotional Eating (13 items), and External Eating (10 items)^17^. All items are rated on a 5-point scale with responses that range from 1 (‘Never’) to 5 (‘Very Often’). Scores for each subscale were calculated by adding up the item responses within a subscale and then dividing by the number of questions in that subscale resulting in a final score per subscale. Higher scores indicate greater tendency to display the subscale behavior.

#### State-Trait Anxiety Inventory-Trait (STAI-T)

Trait-based characteristics relating to general anxiety level were assessed using the STAI-T^15^. The STAI is comprised of 20 item that ask participants to rate level of proneness to anxiety (e.g., general states of calmness, security, or inadequacy) through Likert scale responses that range from 1 (‘Almost Never’) to 4 (‘Almost Always’). Scoring was reversed for the anxiety-absent items, which comprised 9 of the 20 questions. Total scores were calculated by adding the responses after reverse-scoring. The range of scores is from 20-80, with higher scores indicating greater general anxiety.

#### Perceived Stress Scale (PSS)

Participants completed the PSS as part of the post-visit questionnaire packet. The PSS is a 10-item questionnaire that assesses how often respondents have felt a particular way in relation to stressful situations within the past month (e.g., ‘In the last month, how often have you been upset because of something that happened unexpectedly?’)^16^. Each item is rated on a 5-point Likert-scale, with 0 being never and 4 being very often. Six of the items indicating last of perceived stress were reverse-scored. Total scores were calculated by summing the answers to all the scored and reverse-scored items. Higher scored correspond with higher perceived stress.

##### Statistical Methods

Demographic data, questionnaires, and hormone levels were analyzed using SPSS version 28 (IBM Corp., 2021). The p-value threshold for significance was 0.05. Based on small sample sizes and non-normal distributions, non-parametric statistical tests were employed, and serum cortisol and acylated ghrelin were log2 transformed. Demographics, baseline characteristics, BDI-II, STAI-T, DEBQ, PSS, and breakfast meal values (PVC total, PVC protein, PVC fat, and PVC carbohydrate) were assessed using Fisher’s Exact Tests, χ2, and Mann-Whitney U tests. Within group comparisons in hormone levels and VAS ratings of sadness and tension were assessed using Wilcoxon Signed Rank tests. Between-subjects effects in breakfast intake, cortisol and acylated ghrelin samples, and VAS ratings of sadness and tension were analyzed using Mann-Whitney U tests.

### Bulk RNA-seq Analysis

Total RNA was sent to Genewiz (Azenta Life Sciences, Burlington, MA) for bulk-RNAseq. All samples were treated with TURBO DNase (Thermo Fisher Scientific, Waltham, MA) to remove residual DNA contaminants. Ribosomal RNA and globin RNA was depleted using QIAseq FastSelect−rRNA HMR and −Globin kit (Qiagen, Germantown, MD). RNA sequencing libraries were constructed with the NEBNext Ultra II RNA Library Preparation Kit for Illumina. The sequencing libraries were multiplexed and clustered on seven lanes of a flowcell. After clustering, the flowcell was loaded on the Illumina HiSeq 4000 instrument according to manufacturer’s instructions. The samples were sequenced using a 2×150 Pair-End (PE) configuration. Image analysis and base calling were conducted by the HiSeq Control Software (HCS). Raw sequence data (.bcl files) generated from Illumina HiSeq was converted into FASTQ files and de-multiplexed using Illumina’s bcl2fastq 2.17 software. One mismatch was allowed for index sequence identification. The obtained RNAseq data was processed through the bulk RNA-seq pipeline in bcbio (v1.2.8)^19^. Reads were aligned using STAR against transcriptome references of the human genome (GRCh38 Ensembl release 94)^20^ and quantified with Salmon^21^. Two samples were excluded in further analysis due to poor sequencing quality.

### Differential Gene Expression Analysis

Differential gene expression analysis was performed using limma (v3.46.0)^22^. Counts were filtered using limma::filterByExpr with default settings. Counts from both time points were normalized with limma::voom and repeated measures from same individual were accounted for using limma::duplicateCorrelation. To determine which covariates to include in the final analysis, association tests were performed between all known variables (group [Hyperphagic MDD, Hypophagic MDD], visit [Stress, No-Stress], sequencing quality metrics [percent_gc, r_rna, r_rna_rate, intergenic_rate, intronic_rate, exonic_rate, duplication_rate_of_mapped, average_insert_size, total_reads, mapped_reads, mapped_paired_reads, duplicates_pct], individual, sex, age, BMI, visit, breakfast intake [bfast_pvc_TOT, bfast_pvcPROT, bfast_pvcFAT, bfast_pvcCARB], RIN, RNA_extraction_batch) and the 12 calculated principal components (PCs) for each timepoint separately. Some sequencing metrics, RIN, RNA extraction batch, and breakfast intake tested, did not show a significant association with the top PCs (**Supplemental Figure 1**). However, sequencing quality related metrics (intergenic_rate, intronic_rate, exonic_rate, duplication_rate_of_mapped, average_insert_size, and duplicates_pct) showed a significant association with the variation in our data as captured in the top PC and therefore exonic rate was included in the downstream models (p_Bonferroni_ < 0.05). Other sequencing related metrics were not included due to their correlation with exonic rate to prevent collinearity in the model.

At baseline T0, the gene expression differences between Hyperphagic MDD and Hypophagic MDD groups were compared using a linear mixed model controlling for age, BMI, sex, calories of breakfast intake, sequencing quality, and repeated measures. At T105, the gene expression differences between Hyperphagic MDD and Hypophagic MDD groups under stress were compared using a linear mixed model with group and visit interaction terms controlling for age, BMI, sex, calories of breakfast intake, sequencing quality, and repeated measures. Nominal p-values from both models were adjusted for inflation with bacon (v1.18.0)^23^ and differentially expressed genes were considered significant at p_FDR_<0.05.

Normalized counts of all significant genes identified at both time points were tested for correlation with corresponding cortisol and ghrelin levels, breakfast PVC values and mood ratings in linear regression models controlling for sex and repeated measures. Normalized counts at T0 were then averaged between visits to test for association with corresponding DEBQ subscale scores in linear regression models controlling for sex.

### Enrichment Analysis

Differentially expressed genes (DEGs) identified at nominal significance level were used in the enrichment analyses. Overrepresentation tests were used to determine the enrichment of Gene Ontology (GO) terms^24,25^. All genes were ranked based on log fold change and used as input for Gene Set Enrichment Analysis (GSEA) to assess the level of enrichment of 161 inflammatory pathways identified by de Kluiver et al^10^ that are available from the Human MSigDB Gene Set^26,27^. The pathway “Reactome RIGI MDA5 mediated induction of IFN alpha beta pathway” was not found in the Human MSigDB Gene Sets database and excluded in the GSEA analysis.

### Data and Code Availability

All RNA-seq statistical analyses were performed in R software version 4.0.2. Bulk RNA-seq data can be accessed through GEO Accession GSE231347. Code used in the analysis has been deposited on Github: https://github.com/klengellab/HypoHyperMDD.

## Results

### Demographic and Trait Characteristics

Direct comparison between Hyperphagic MDD and Hypophagic MDD revealed that groups did not differ in demographic characteristics (sex [*p*=1.000], age [z= -0.545, *p*=0.595], BMI [z=-0.072, *p*=0.972], race [*p*=0.118], ethnicity [*p*=0.618]), or trait-level anxiety/stress-related characteristics (STAI-T [z=-1.56, *p*=0.121], PSS [z=-1.38, *p*=0.185]) (**Table 1**). Groups differed in self-reported depression symptom severity (BDI-II), with Hyperphagic MDD exhibiting more severe depression symptoms (z=2.24, *p*=0.025). There was a significant difference between groups in Emotional Eating (z=-2.62, *p*=0.008) and External Eating (z=-2.04, *p*=0.045) subscales of the DEBQ, with Hyperphagic MDD exhibiting elevated levels of emotional and external eating behavior compared to Hypophagic MDD. There were no differences between groups in the Restrained Eating subscale (z=-1.16, *p*=0.268). For breakfast meal intake, Hyperphagic MDD displayed higher PVC kcals from protein (z=2.14, p=0.030) compared with Hypophagic MDD. Groups did not differ in PVC total kcals (z=1.85, p=0.064), PVC kcals from fat (z=1.78, p=0.076), or PVC kcals from carbohydrates (z=1.41, p=0.161).

**Table 1.**
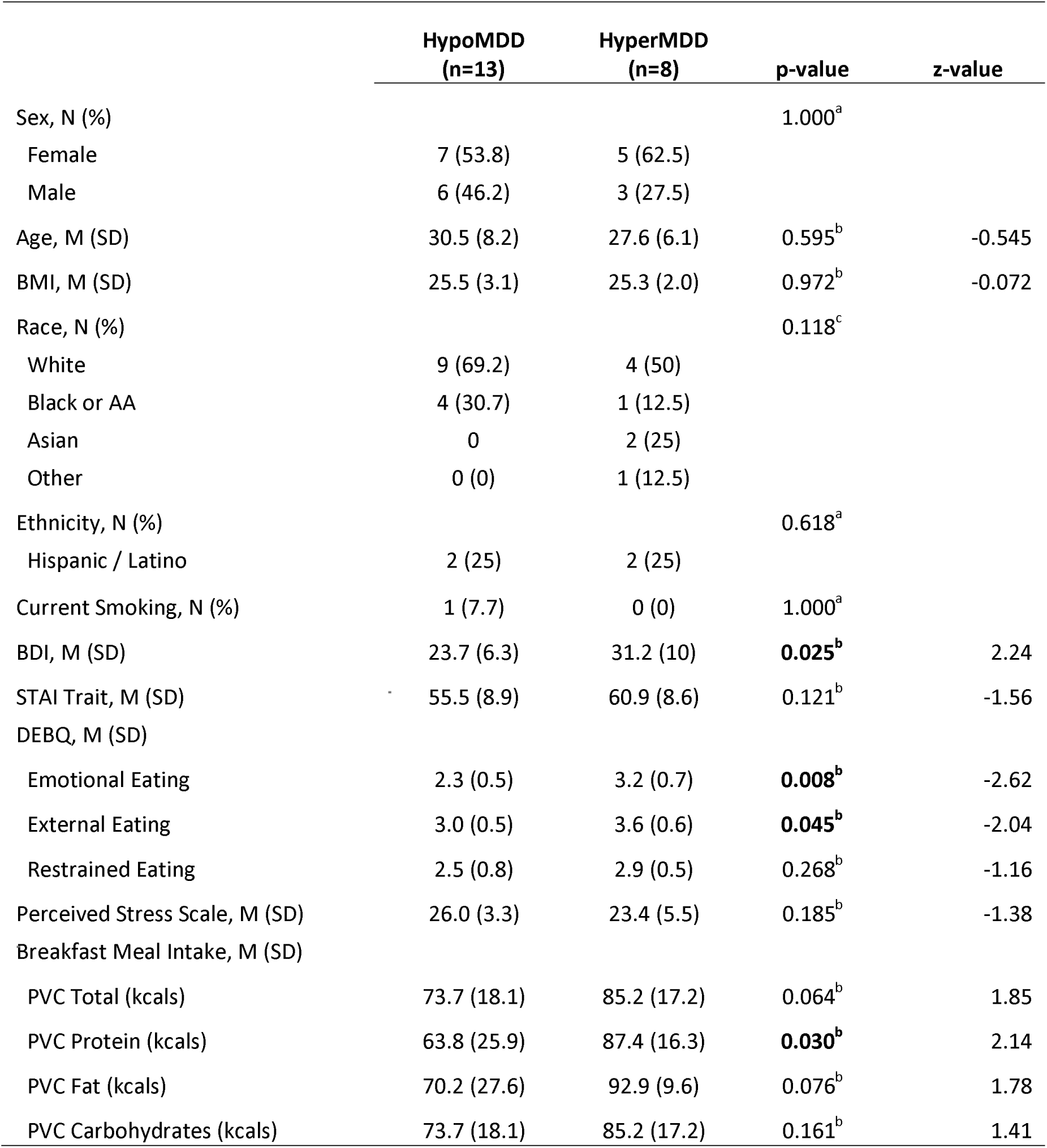
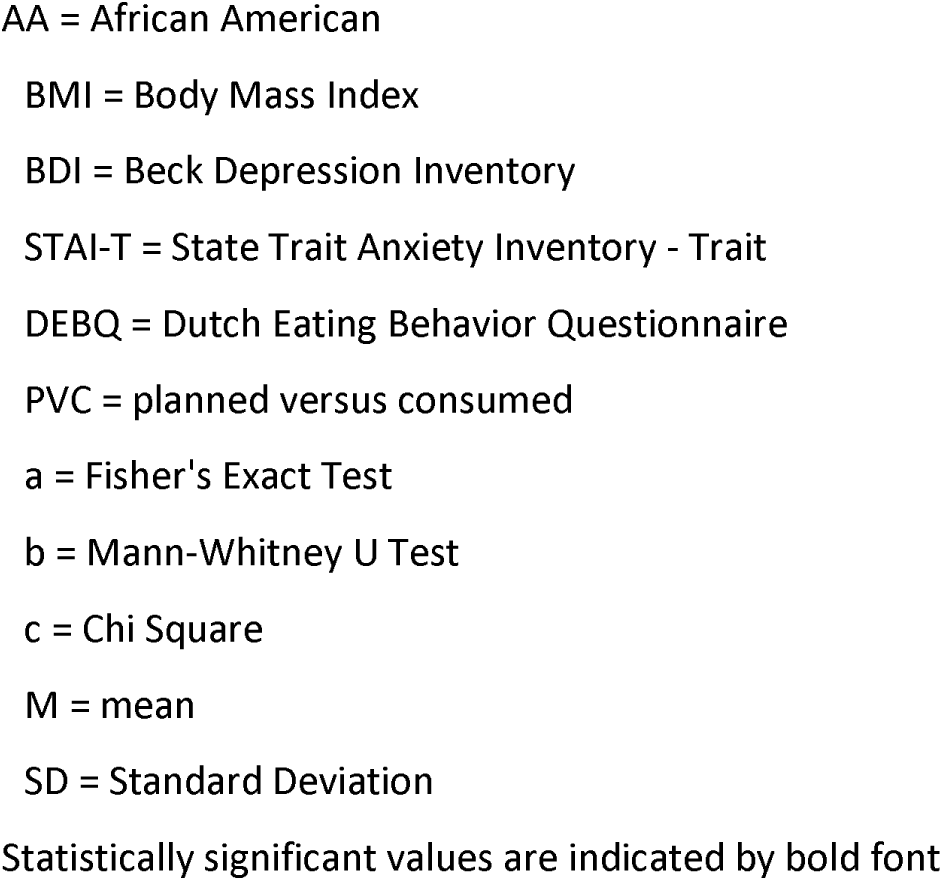
Demographic, Trait, and Baseline Variables.

|  | HypoMDD<br>(n=13) | HyperMDD<br>(n=8) | p-value | z-value |
| --- | --- | --- | --- | --- |
| Sex, N (%) |  |  | 1.000<sup>a</sup> |  |
| Female | 7 (53.8) | 5 (62.5) |  |  |
| Male | 6 (46.2) | 3 (27.5) |  |  |
| Age, M (SD) | 30.5 (8.2) | 27.6 (6.1) | 0.595<sup>b</sup> | -0.545 |
| BMI, M (SD) | 25.5 (3.1) | 25.3 (2.0) | 0.972<sup>b</sup> | -0.072 |
| Race, N (%) |  |  | 0.118<sup>c</sup> |  |
| White | 9 (69.2) | 4 (50) |  |  |
| Black or AA | 4 (30.7) | 1 (12.5) |  |  |
| Asian | 0 | 2 (25) |  |  |
| Other | 0 (0) | 1 (12.5) |  |  |
| Ethnicity, N (%) |  |  | 0.618<sup>a</sup> |  |
| Hispanic / Latino | 2 (25) | 2 (25) |  |  |
| Current Smoking, N (%) | 1 (7.7) | 0 (0) | 1.000<sup>a</sup> |  |
| BDI, M (SD) | 23.7 (6.3) | 31.2 (10) | **0.025<sup>b</sup>** | 2.24 |
| STAI Trait, M (SD) | 55.5 (8.9) | 60.9 (8.6) | 0.121<sup>b</sup> | -1.56 |
| DEBQ, M (SD) |  |  |  |  |
| Emotional Eating | 2.3 (0.5) | 3.2 (0.7) | **0.008<sup>b</sup>** | -2.62 |
| External Eating | 3.0 (0.5) | 3.6 (0.6) | **0.045<sup>b</sup>** | -2.04 |
| Restrained Eating | 2.5 (0.8) | 2.9 (0.5) | 0.268<sup>b</sup> | -1.16 |
| Perceived Stress Scale, M (SD) | 26.0 (3.3) | 23.4 (5.5) | 0.185<sup>b</sup> | -1.38 |
| Breakfast Meal Intake, M (SD) |  |  |  |  |
| PVC Total (kcal) | 73.7 (18.1) | 85.2 (17.2) | 0.064<sup>b</sup> | 1.85 |
| PVC Protein (kcal) | 63.8 (25.9) | 87.4 (16.3) | **0.030<sup>b</sup>** | 2.14 |
| PVC Fat (kcal) | 70.2 (27.6) | 92.9 (9.6) | 0.076<sup>b</sup> | 1.78 |
| PVC Carbohydrates (kcal) | 73.7 (18.1) | 85.2 (17.2) | 0.161<sup>b</sup> | 1.41 |

AA = African American BMI = Body Mass Index BDI = Beck Depression Inventory STAI-T = State Trait Anxiety Inventory - Trait DEBQ = Dutch Eating Behavior Questionnaire PVC = planned versus consumed a = Fisher's Exact Test b = Mann-Whitney U Test c = Chi Square M = mean SD = Standard Deviation Statistically significant values are indicated by bold font

### Acute effect of MAST on cortisol levels

To assess the acute physiological response to stress, cortisol was measured and analyzed at T0 and T20. At the No-Stress visit, there were no significant acute changes in log2 cortisol from T0 to T20 for either the Hyperphagic MDD group (z=0.338, p=0.735) or Hypophagic MDD group (z=1.818, p=0.069). At the Stress visit, an increase in log2 cortisol from T0 to T20 was observed for the Hyperphagic MDD group (z=2.197, p=0.028), but not for the Hypophagic MDD group (z=0.070, p=0.944)

### Cortisol and Ghrelin Responses

Wilcoxon Signed Rank tests were used to assess the within-group effect of time in each group at the Stress and No-Stress visits (**Table 2**). Log2 cortisol was significantly higher at T0 vs T105 in both Hypophagic MDD (z=-2.71, *p*=0.006) and Hyperphagic MDD (z=-2.24, *p*=0.025) at the No-Stress Visit. During the Stress visit, log2 cortisol was significantly elevated at T0 vs. T105 in Hypophagic MDD only (z=-2.06, *p*=0.039), with no significant change in Hyperphagic MDD (*z*=1.35, *p*=0.176). There was not a within-subject effect of time on log2 ghrelin in either group or at either visit (No-Stress Hypophagic MDD [z=-1.78, *p*=0.075], No-Stress Hyperphagic MDD [z=-0.42, *p*=0.674], Stress Hypophagic MDD [z=-1.22, *p*=0.221], Stress Hyperphagic MDD [z=-1.26, *p*=0.208]).

**Table 2.**
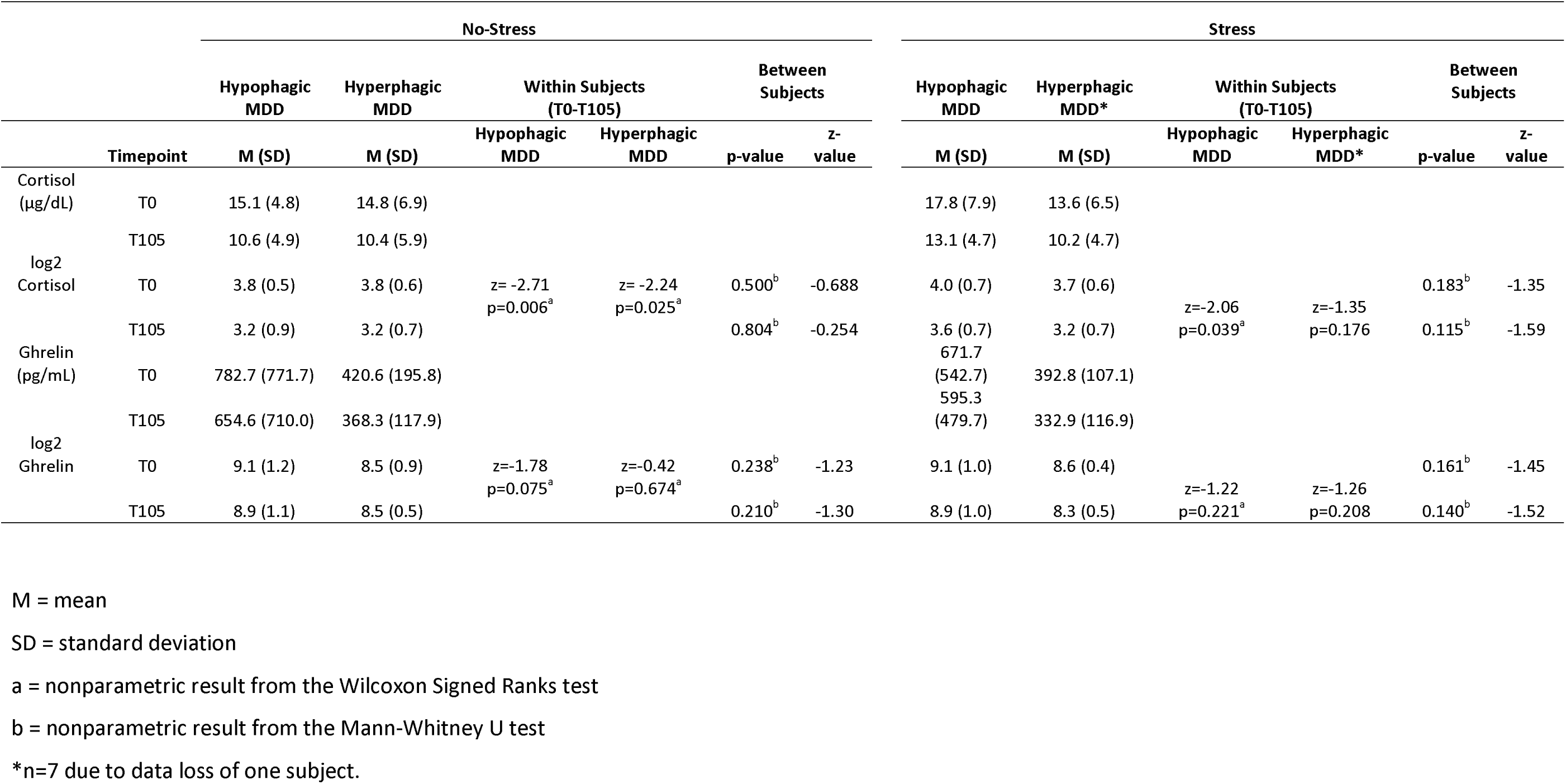
Hormone responses.

Mann-Whitney U tests were used to compare serum levels of cortisol and acylated ghrelin between groups at the Stress and No-Stress visits (**Table 2**). There were no significant differences found in log2 cortisol at T0, log2 cortisol at T105, log2 acylated ghrelin at T0, or log2 acylated ghrelin at T105 between Hyperphagic MDD and Hypophagic MDD at either the Stress or No-Stress visits.

### Mood Ratings

Mood was assessed by comparing ratings of tension and sadness between Hyperphagic MDD and Hypophagic MDD at T0 and T105 for each visit (**Table 3**). Within-subject analyses revealed an effect of time on ratings of tension in Hyperphagic MDD at the Stress visit (z=2.10, p=0.035). There were no other within-subjects effects of time on sadness (No-Stress Hypophagic MDD [z=-1.07, p=0.286], No-Stress Hyperphagic MDD [z=0.676, p=0.499], Stress Hypophagic MDD [z=-1.16, p=0.248], Stress Hyperphagic MDD [z=-.508, p=0.611]) or tension (No-Stress Hypophagic MDD [z=0.711, p=0.477], No-Stress Hyperphagic MDD [z=1.47, p=0.141], Stress Hypophagic [z=0.157, p=0.875]). Between-subjects analyses revealed that Hyperphagic MDD reported feeling significantly more tense than Hypophagic MDD at T105 in both the Stress (z=-1.98, *p*=0.047) and No-Stress Visit (z=-2.051, *p*=0.045). There were no additional differences found between groups in ratings of sadness or tension at either visit.

**Table 3.**
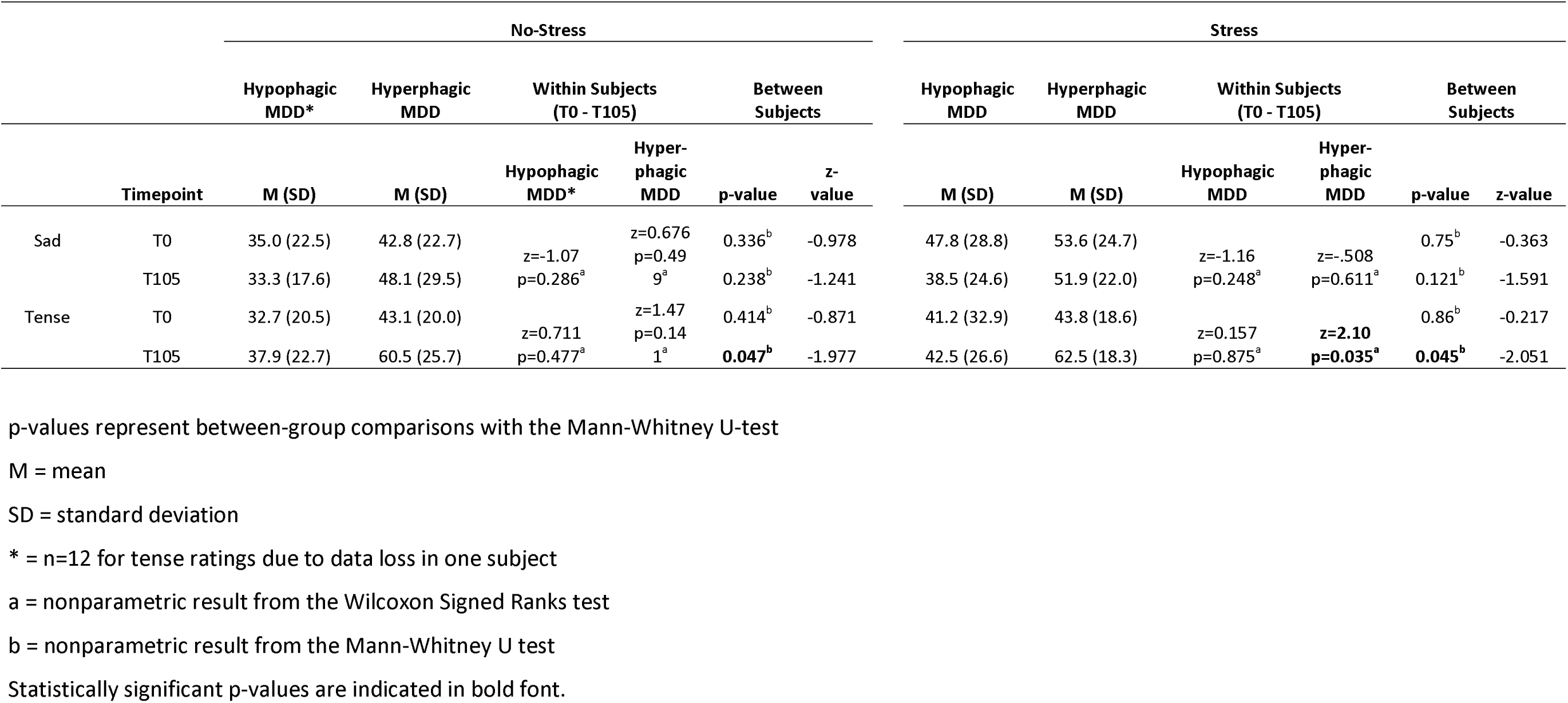
Mood Ratings.

### Baseline Transcriptomic Differences

At baseline T0 when controlling for age, BMI, sex, calories of breakfast intake, sequencing quality, and repeated measures, the expression of transcriptional adaptor 2-beta (*TADA2B*) gene was significantly increased in the Hyperphagic MDD group in comparison to the Hypophagic MDD group after controlling for multiple testing (logFC= 0.22, *p_FDR_* = 0.013) (**Figure 2A**, QQ-plot in **Supplemental Figure 2**, full list of nominal significant DEGs in **Supplemental Table 1**). The expression of *TADA2B* was positively associated with the DEBQ external eating scores (ß = 0.107, *p* = 0.024) and PVC fat (ß = 0.0017, *p* = 0.025) (**Table 4**). However, no significant associations of *TADA2B* expression with log2 cortisol, log2 ghrelin, mood ratings, other DEBQ subscales or breakfast intake were detected.

**Figure 2.**
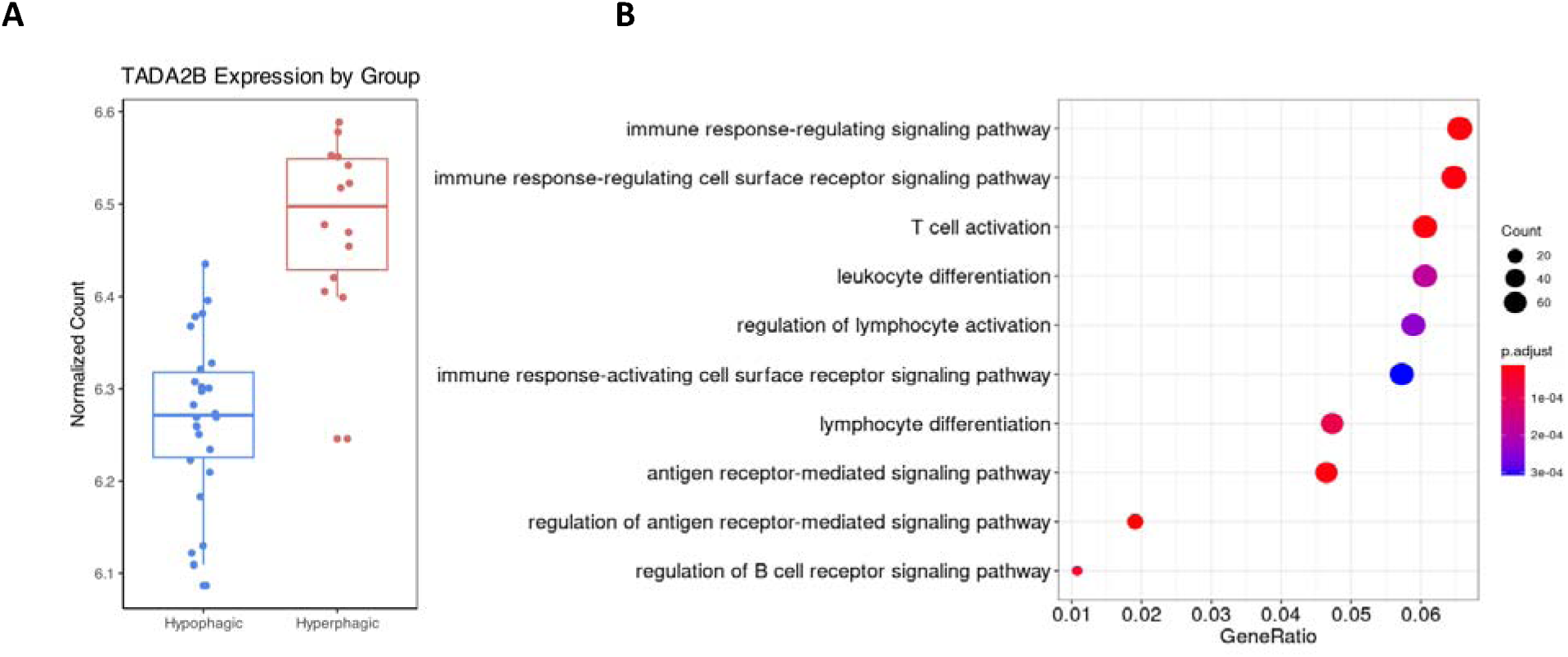
Baseline transcriptional differences at baseline T0. (**A**) TADA2B was found to be differentially expressed between Hyperphagic and Hypophagic MDD at baseline. The expression of TADA2B is elevated in Hyperphagic MDD in comparison to Hypophagic MDD. (**B**) Gene Ontologies related to immune signaling pathways were found to be most significantly enriched using nominally significant differentially expressed genes (p_nominal_ < 0.05).

**Table 4.**
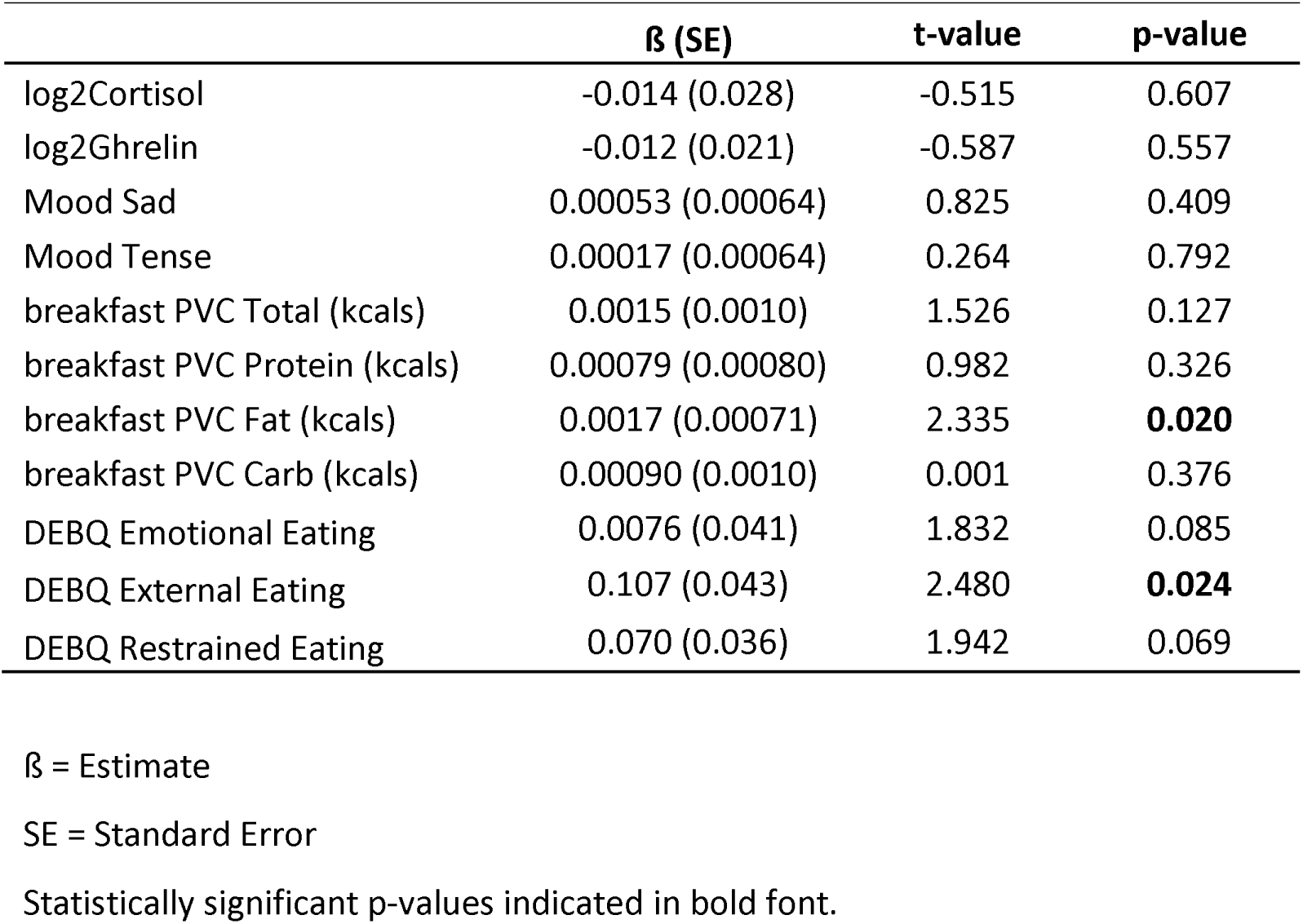
Association between TADA2B expression and other measurements.

|  | **β (SE)** | **t-value** | **p-value** |
| --- | --- | --- | --- |
| log2Cortisol | -0.014 (0.028) | -0.515 | 0.607 |
| log2Ghrelin | -0.012 (0.021) | -0.587 | 0.557 |
| Mood Sad | 0.00053 (0.00064) | 0.825 | 0.409 |
| Mood Tense | 0.00017 (0.00064) | 0.264 | 0.792 |
| breakfast PVC Total (kcal) | 0.0015 (0.0010) | 1.526 | 0.127 |
| breakfast PVC Protein (kcal) | 0.00079 (0.00080) | 0.982 | 0.326 |
| breakfast PVC Fat (kcal) | 0.0017 (0.00071) | 2.335 | **0.020** |
| breakfast PVC Carb (kcal) | 0.00090 (0.0010) | 0.001 | 0.376 |
| DEBQ Emotional Eating | 0.0076 (0.041) | 1.832 | 0.085 |
| DEBQ External Eating | 0.107 (0.043) | 2.480 | **0.024** |
| DEBQ Restrained Eating | 0.070 (0.036) | 1.942 | 0.069 |

β = Estimate SE = Standard Error Statistically significant p-values indicated in bold font.

72 significantly enriched Gene Ontologies (GO) terms were found based on the nominal significant differentially expressed genes (*p_nominal_*<0.05, n=1556). The top 10 GO terms fall exclusively under the biological processes sub-category relating to inflammation pathways, including the regulation of antigen receptor-mediated signaling pathway, immune response-regulating signaling pathway, and T cell activation pathway (*p_FDR_* < 0.05, q-value < 0.05) (**Figure 2B, Supplemental Table 2**).

De Kluiver et al^10^ previously described an enrichment of inflammatory pathways in Hyperphagic MDD and -to a lesser extent-Hypophagic MDD relative to healthy individuals. While replicating findings from de Kluiver et al, 8 previously described inflammation-related pathways (*p_FDR_* < 0.1, q-value < 0.1) were significantly enriched in the Hyperphagic versus Hypophagic group comparison in our study (**Supplemental Table 5**). Among these enriched inflammatory pathways, KEGG T cell receptor signaling was also found to be significantly enriched in the combined MDD and Hyperphagic MDD groups in the de Kluiver et al study (*p_FDR_* < 0.05).

### Differential Gene Expression Response to the Maastricht Acute Stress Task

A defined stressor may unmask additional stress-responsive differences on the level of RNA expression between Hyperphagic MDD and Hypophagic MDD. In fact, RNAseq profiling 85 minutes after the MAST revealed a subtype-specific differential response of Coiled-Coil Domain Containing 196 (*CCDC196*) (logFC = 2.10, *p_FDR_* = 0.00063) and Spermatogenesis Associated 33 (*SPATA33*) (logFC = -0.87, *p_FDR_* = 0.0495) significantly differentially expressed between Hyperphagic MDD and Hypophagic MDD (**Figure 3A-B**, QQ-plot in **Supplemental Figure 3**, full list of nominal significant DEGs in **Supplemental Table 3**). The expression of *CCDC196* decreased in the Hypophagic MDD group after stress, while increasing in the Hyperphagic MDD group after stress. *CCDC196* expression was also positively associated with PVC protein consumed during breakfast (ß = 0.0075, *p* = 0.007). Conversely, the expression of *SPATA33* increased in the Hypophagic MDD and decreased in the Hyperphagic MDD group after stress. Beyond group differences, the expression of *SPATA33* was negatively associated with stress-induced cortisol changes (ß = -0.54, *p* = 0.0050) and positively associated with PVC protein consumed during breakfast (ß = 0.012, *p* = 0.049) (**Table 5**).

**Figure 3.**
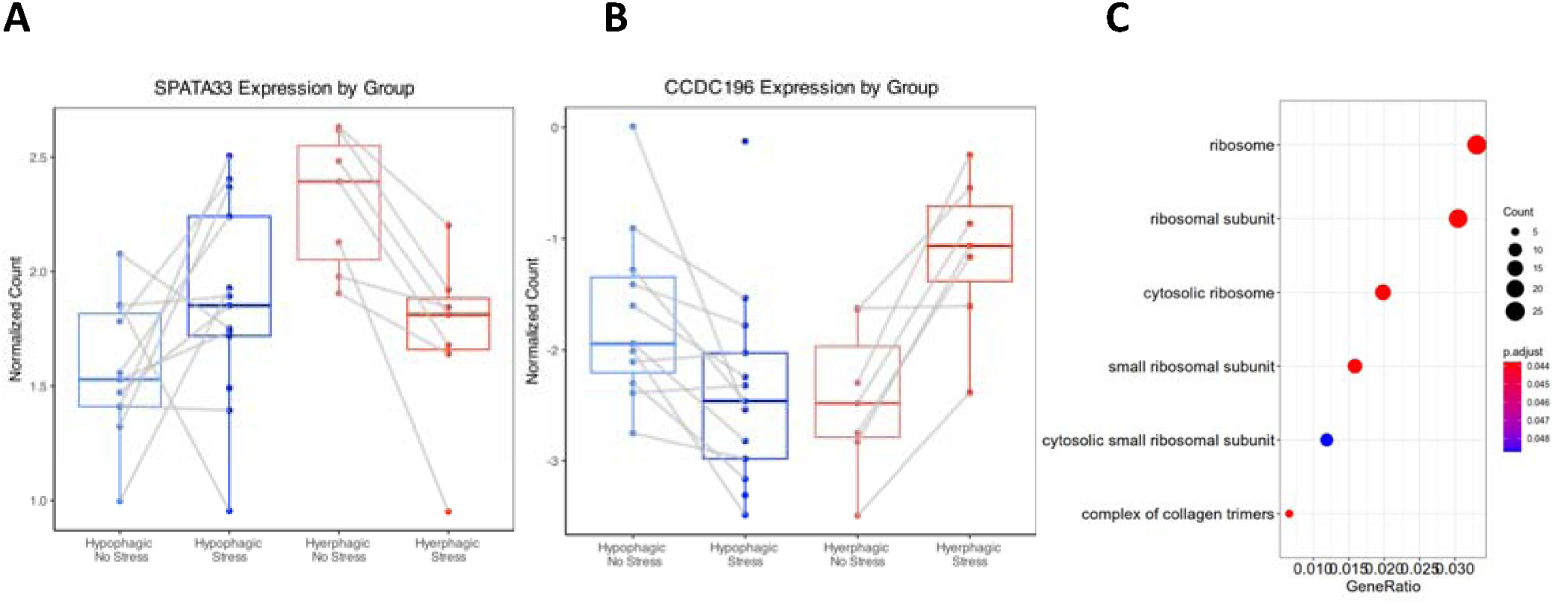
Differential transcriptional changes in response to stress at T105. (**A**) SPATA33 was found to be differentially expressed between Hyperphagic and Hypophagic MDD after MAST. The expression of SPATA33 was decreased in Hyperphagic MDD while increasing to Hypophagic MDD. (**B**) CCDC196 was also found to be differentially expressed in response to stress. Conversely to the expression of SPATA33, the expression of CCDC196 increased in Hyperphagic MDD and decreased in Hypophagic MDD. (**C**) 7 Gene Ontologies were found to be significantly enriched using nominally significant differentially expressed genes (p_nominal_ < 0.05).

**Table 5.** Association between stress-induced transcripts expression and other measurements.

|  | CCDC196 |  |  | SPATA33 |  |  |
| --- | --- | --- | --- | --- | --- | --- |
| | $\beta$ (SE) | t-value | p-value | $\beta$ (SE) | t-value | p-value |
| log2Cortisol | 0.028 (0.099) | 0.279 | 0.780 | -0.54 (0.18) | -3.000 | **0.0027** |
| log2Ghrelin | -0.15 (0.077) | -1.903 | 0.057 | -0.12 (0.16) | -0.759 | 0.448 |
| Mood Sad | 0.0032 (0.0032) | 1.009 | 0.313 | -0.0011<br>(0.0067) | -0.169 | 0.866 |
| Mood Tense | 0.0059 (0.0031) | 1.920 | 0.055 | 0.0038<br>(0.0066) | 0.587 | 0.557 |
| breakfast PVC Total<br>(kcal) | 0.0069 (0.0038) | 1.800 | 0.072 | 0.011<br>(0.0080) | 1.336 | 0.182 |
| breakfast PVC<br>Protein (kcal) | 0.0075 (0.0028) | 2.695 | **0.007** | 0.012<br>(0.0059) | 2.035 | **0.042** |
| breakfast PVC Fat<br>(kcal) | 0.0036 (0.0031) | 1.174 | 0.240 | 0.0064<br>(0.0063) | 1.015 | 0.310 |
| breakfast PVC Carb<br>(kcal) | 0.0051 (0.0038) | 1.331 | 0.183 | 0.0072<br>(0.0079) | 0.906 | 0.365 |

Similar to baseline, Gene Ontology enrichment analyses on nominal significant DEGs (*p_nominal_*<0.05, n = 1296) suggested 6 significantly enriched GO terms that are all ribosomal activity relevant cellular component sub-ontologies (*p_FDR_* < 0.05, q-value < 0.05) (**Figure 3C, Supplemental Table 4**). The differential stress-induced gene set was not enriched in any previously described inflammatory pathways from de Kluiver et al.

## Discussion

Eating behaviors are important endophenotypes of MDD and despite their rather simple assessment, they remain understudied although appetite and weight are critical determinants of therapy response^28^, medication side effects, and comorbidities^29^. Prior studies including our work provided evidence for distinct neural networks involved in Hyper- and Hypophagic MDD, with hyperresponsivity and connectivity in Hyperphagic MDD in reward circuits and hypofunctioning within interoceptive circuits in Hypophagic MDD. Molecular, genetic, endocrine, and metabolic studies of hyper- and hypophagia in MDD suggest that appetite, weight gain, obesity and treatment response are intertwined at the level of stress-hormone and orexigenic signaling, immune-metabolic changes, and genetic disposition. Only one study by de Kluiver et al so far investigated the biological correlates of Hyper- and Hypophagic MDD at the level of gene expression in peripheral blood using gene expression arrays. Our study aimed to replicate and expand prior work and to perform a deep phenotyping of hyper- and hypophagia in unmedicated MDD patients and to analyze their differential transcriptomic profile changes in response to stress using RNAseq.

In line with the handful of prior reports detailing demographic and clinical characteristics of appetite and weight phenotypes in MDD, this small sample of unmedicated individuals with Hyper- and Hypophagic MDD had similar BMIs and endorsed comparable levels of general anxiety and perceived stress. Notably, however, in contrast to our prior report on women with *remitted* MDD, in whom scores on depressive symptomatology and domains of eating behavior (disinhibition, restraint, hunger) were similar for opposing appetitive subgroups, those with Hyperphagic MDD in the current mixed-sex sample reported elevated severity of depressive symptoms and of emotional and external eating than the Hypophagic MDD group^4^. This suggests that, among those with Hyperphagic MDD, (1) depressive symptoms may be more severe; and (2) eating behavior domains may vary with current state, such that internal and external influences on eating characteristics become more pronounced during depressive episodes.

Moreover, although raw levels of baseline cortisol and ghrelin appeared higher in the Hypophagic MDD group, similar to prior findings^1^, comparison of log-transformed values yielded no significant group differences. This may have been due to small sample sizes in each group attenuating statistical power to detect effects. Finally, we noted that based on within-group comparisons of T0 vs. T105 timepoints, the Hyperphagic MDD exhibited a sustained cortisol response to stress that was absent in the Hypophagic MDD group. This effect appeared to be mirrored in ratings of tension, which remained elevated at T105 in the Hyperphagic MDD group. To our knowledge, this is the first study to employ a stress manipulation to examine mood and hormone changes to psychosocial stress in opposing appetite/weight phenotypes in MDD.

To identify biological correlates related to the divergent appetite-related phenotypes described, we performed an unbiased RNAseq study at baseline and after exposure to a defined stressor. At baseline, after the intake of a standardized breakfast meal (T0), the expression of *TADA2B* (Transcriptional Adaptor 2B) differentiates Hyper- and Hypophagic individuals with MDD with higher expression levels in hyperphagia when controlling for relevant covariates (**Figure 2A**). *TADA2B* is a protein coding gene that participates histone acetylation and chromatin remodeling. It has been described primarily in the context of stem cell biology and transcriptional regulation^30,31^. However, evidence for a role of *TADA2B* in psychiatric disorders is limited to a potential involvement in autism spectrum disorder^32^, and to the best of our knowledge, *TADA2B* has not been described in the context of MDD or eating disorders. In the pathway analysis of the gene expression signature at baseline, we observed significant enrichment of immune-related gene ontologies in Hyperphagic MDD, suggesting a potential differentiation between these sub-phenotypes based on variations in immune signaling. This finding was reinforced by the overlap identified through GSEA against curated immune pathways established by de Kluiver et al., replicating aspects of prior studies and providing further evidence of disrupted immune signaling as a hallmark of divergent appetitive phenotypes in MDD.

As discussed before, HPA axis signaling has been investigated extensively in the context of MDD. Evidence suggests that differences in cortisol levels and HPA axis responsivity may differentiate subgroups of individuals with MDD, particularly in individuals with vegetative symptoms including changes in appetite^33^. We applied the Maastricht Acute Stress Task (MAST) in association with RNAseq profiling to uncover stress-induced transcriptional changes in depressed individuals with Hyper- and Hypophagia. Differential gene expression analysis revealed that *CCDC196* (Coiled-Coil Domain Containing 196) and *SPATA33* (Spermatogenesis Associated 33) are differentially expressed between Hyper- and Hypophagia after the MAST. These two novel stress induced transcripts have not been described in the context of psychiatric diseases. *CCDC196*, also known as Long Intergenic Non-Protein Coding RNA 238 (*LINC00238*), encodes a coiled-coil domain-containing (CCDC) protein. CCDC proteins are expressed in various tissue types, particularly in the human reproductive system, and have been identified in the pathological process of migration, proliferation, and metastasis of cancers^34^. Lower expression of *LINC00238* has been associated with liver cancer and the overexpression of *LINC00238* has been confirmed to act as a tumor suppressor through miR-522 in liver cancer tissues^35,36^. *SPATA33* is mainly expressed in the human reproductive system and participates in spermatogenesis^37,38^. It has also been associated with melanoma risk^39^ and although *SPATA33* has not been extensively studied in neuropsychiatric disease, Liu et al provided evidence on the expression of hypothalamic *SPATA33* changes in response to different calorie restriction interventions in mice^40^. This finding indicates a potential role of *SPATA33* in metabolic regulation of hunger. However, further studies are needed to investigate the mechanism of these specific genes and establish additional evidence for their potential role as biomarkers of Hypophagic and Hyperphagic MDD.

There are several limitations in the current study. Given the small sample sizes, additional data is required to examine responsivity to psychosocial stress in larger samples of individuals with Hyper- and Hypophagic MDD. Additionally, despite stringent statistical methods of all gene expression analyses, an independent replication in a larger cohort is warranted. In addition, our sample consisted primarily of individuals with Caucasian decent thus future studies need to include a more diverse study population. Even though limitations may narrow the generalizability of this study, we replicate prior work and importantly, provide additional phenotypic and molecular data on unmedicated individuals with Hyperphagic and Hypophagic MDD.

## Author contributions

TK and LMH designed research; TK, LMH, SD, JH conducted research; SD, TK, JH, and LMH analyzed data; SD and JH wrote the first draft of the manuscript and LMH and TK made substantive revisions; TK and LMH had primary responsibility for final content. All authors read and approved the final manuscript.

## Acknowledgements

This study was supported by grants from the NIDDK (R01 DK104772) and from the Connors Center for Women’s Health and Gender Biology at Brigham and Women’s Hospital. We would like to thank Sarah Boukezzi for her contributions to initial aspects of study design, Harlyn Aizley and Florina Haimovici for clinical assessments, Rose Chang for assistance with project coordination. We are grateful to Jill Goldstein, Diego Pizzagalli, and Daniel Dillon with their contributions to R01 DK104772. Work in the Klengel Lab is supported by NICHD R01 HD102974, the Connor Group Kids and Community Partners, and NIA R01 AG070704.

## Supplementary Data in Separate Files

**Supplemental Figure 1A**

PCA association plot of covariates and calculated PCs at baseline.

**Supplemental Figure 1B**

PCA association plot of covariates and calculated PCs after stress task.

**Supplemental Figure 2**

QQ plots with lambda values for baseline transcriptomic difference comparison.

**Supplemental Figure 3**

QQ plots with lambda values for stress-induced transcriptomic difference comparison.

**Supplement Table 1**

Differentially expressed genes at T0

**Supplement Table 2**

Differentially expressed genes at T105

**Supplement Table 3**

Enriched GO terms at T0

**Supplement Table 4**

Enriched GO terms at T105

**Supplement Table 5**

Enriched de Kluvier et al. pathways at T0

