## SupplementalMaterial for "Divergent transcriptomic profiles in depressed individuals with hyper- and hypophagia implicating inflammatory status": SupplementalFigures.pptx

### Slide 1
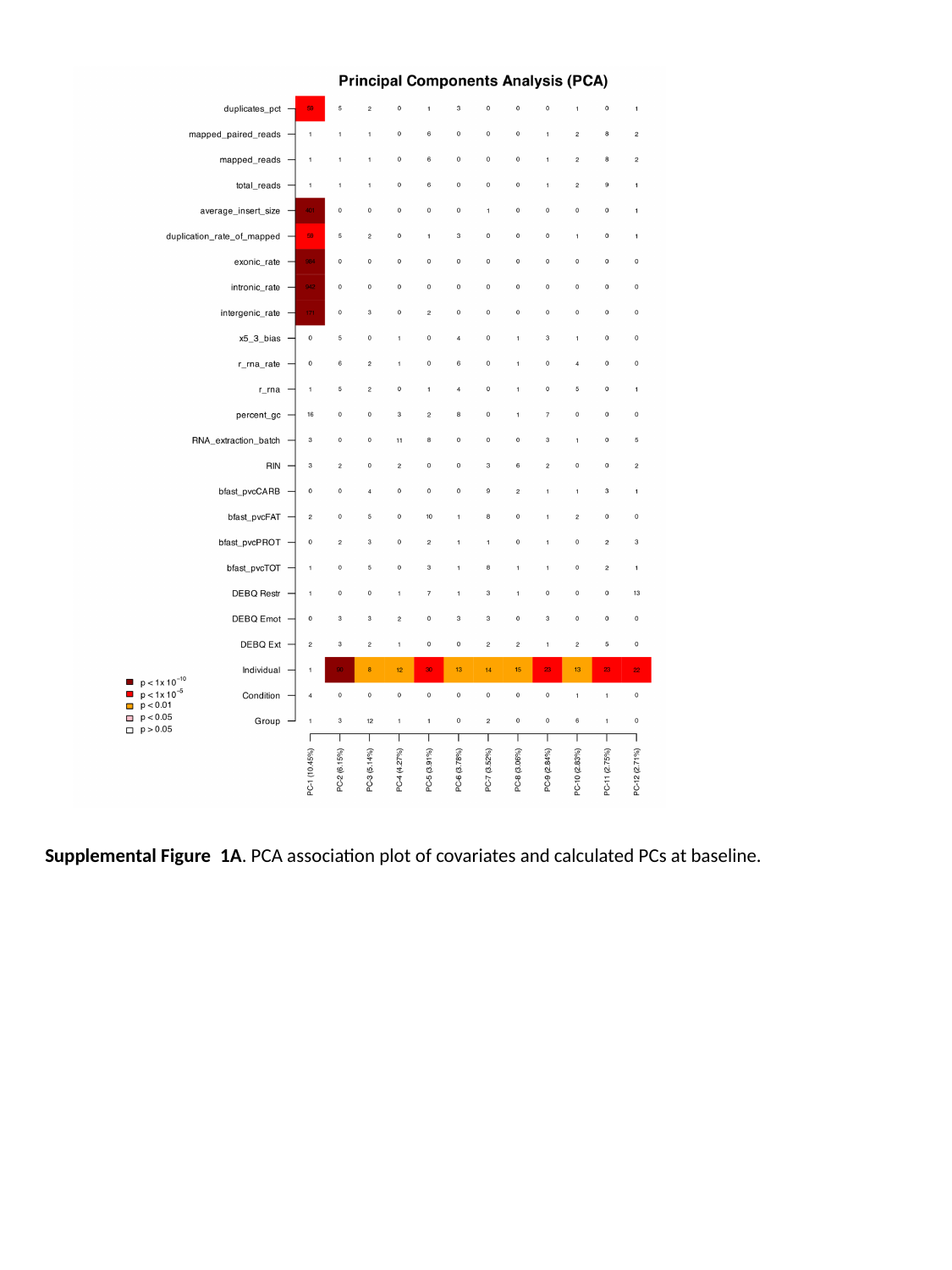

Supplemental Figure  1A. PCA association plot of covariates and calculated PCs at baseline.

### Slide 2
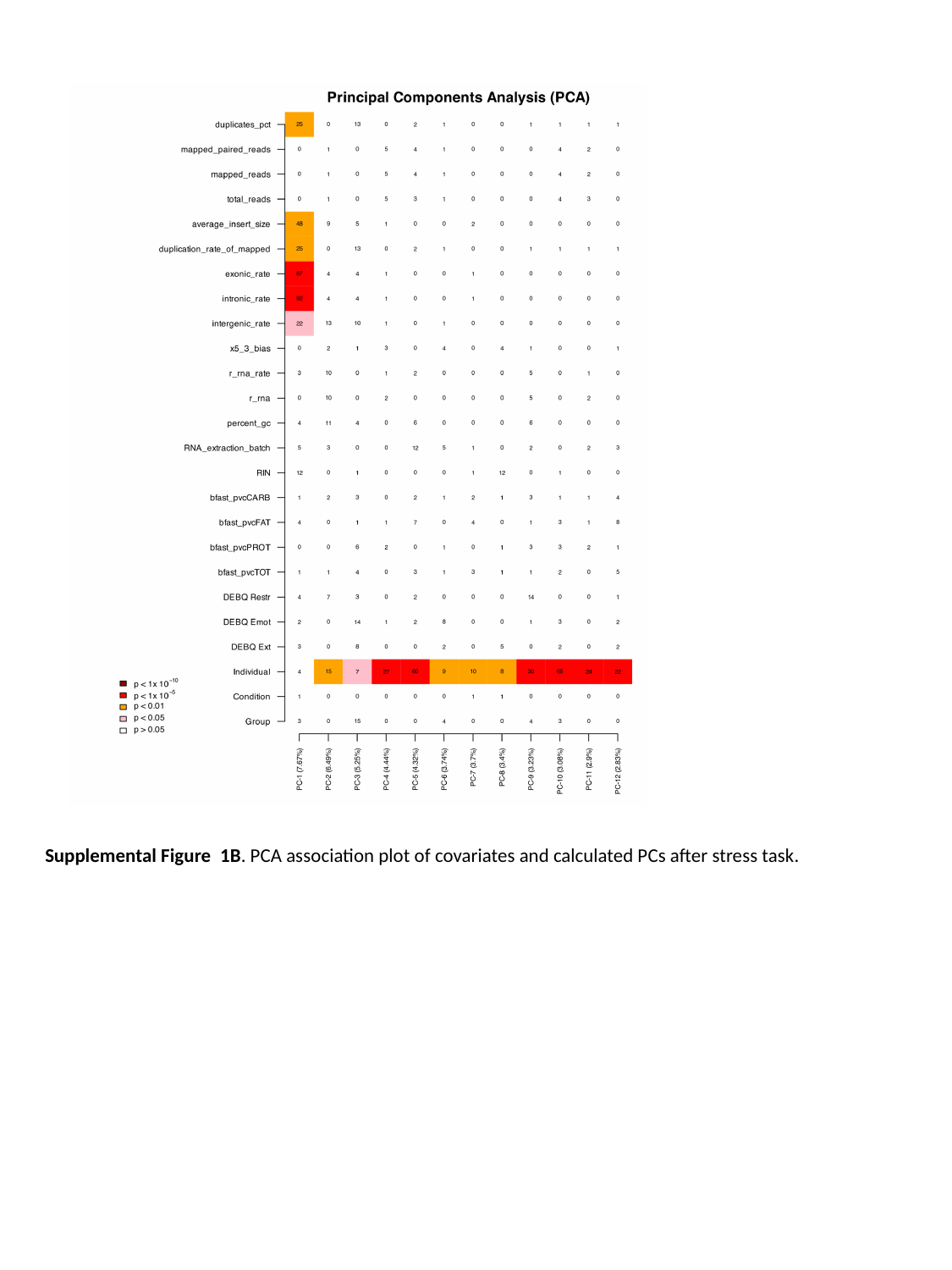

Supplemental Figure  1B. PCA association plot of covariates and calculated PCs after stress task.

### Slide 3
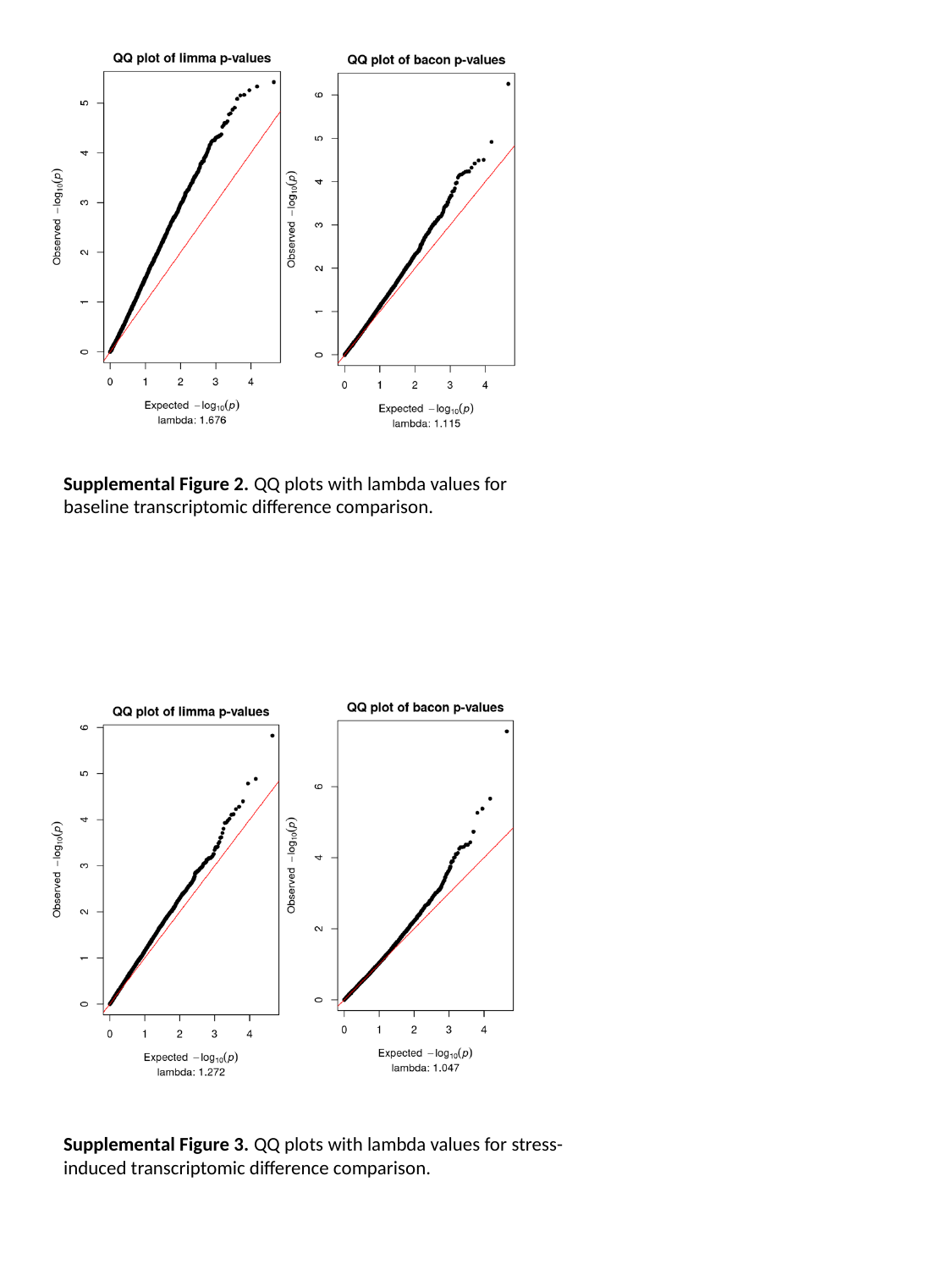

Supplemental Figure 2. QQ plots with lambda values for baseline transcriptomic difference comparison.
Supplemental Figure 3. QQ plots with lambda values for stress-induced transcriptomic difference comparison.
